# Philosophy Spirituality and Recovery of Mental Health Nursing: Literatur Review

**DOI:** 10.1101/2022.10.20.22281082

**Authors:** Sawab, Moses Glorino Rumambo Pandin, Ah Yusuf

## Abstract

**Background:** The recurrence rate and quality of life for mental disorders are still quite low. The phenomenon of mental recovery is still focused on clinical recovery, which emphasizes drug therapy. On the other hand, empowering the patient to be able to control themselves to get a meaningful life from their illness becomes a challenge. Mental health nursing focuses on the well-being of individuals so that they have a good quality of life. Spirituality-based psychological management helps patients with mental disorders to have self-awareness, self-efficacy, self-empowerment and to create meaningful life as an important intervention in addition to drug administration. This literature review aims to explore the influence of spirituality on mental health recovery.

**Methods:** This study used a literature review design with four databases: Scopus, Science Direct, ProQuest, and SpringerLink. There were 12 reviewed articles published in 2019-2022. Keywords used are 1) spirituality or religiosity AND recovery AND mental illness. 2) spirituality or religiosity AND recovery AND severe mental disorder. Protocol and literature review evaluation use PRISMA.

**Results:** Spirituality helps mental disorder patients to build self-confidence, control themselves, find strength and build hope to find the meaning of their life. The ability to control themselves and find the meaning of their life becomes psychological well-being that will be able to improve the quality of life.

**Conclusion:** Recovery from mental disorders is complex and multifactorial; therefore, psychological interventions are constantly being developed. Psychological interventions using spirituality by involving sufferers, nurses, families, religious leaders, and other health teams need to be developed.

## 1. Background

Mental health nursing is a specialized nursing practice committed to promoting mental health through the assessment, diagnosis, and treatment of behavioral problems, as well as lifelong comorbidities. Mental health nurses provide comprehensive and individual-centered care in the nursing process. An important component of mental health nursing practice is health promotion and well-being improvement through the identification of mental health problems, prevention, treatment of mental health problems, and treatment of people with mental disorders, including medication use disorders (Nurses Association, 2014). The global trend toward mental health interventions continues to undergo a transformation from service-oriented biomedical approaches to *recovery* approaches (Evan K. et al., (2020). A conceptual approach model in the recovery of mental health has been widely developed. Watson, in Human Caring Model, explained that the relationship between the individual and the nurse could improve the individual’s self-recovery and develop awareness. Three important things in this model are interpersonal relationships, caring attitudes, and willingness to help the recovery process (Çam, 2017). Recovery Alliance Theory explains that during recovery, individuals are able to make their own choices, and they have the potential to increase their self-awareness, as well as their interaction with themselves and others. This model allows psychiatric patients and nurses to develop an understanding of others (Jubb & Shanley, 2007).

Mental health recovery, according to the Substance Abuse and Mental Health Service Administration (SAMSHA) is the journey and individuals’ transformation with mental health problems in order to live meaningfully with limitations due to their illness. The fundamental components in the concept of recovery are individual, self-empowering, able to control themselves, using their own strengths, holistic support, respect, and building hope (Whitley R, 2010). Recovery is directed at empowering individuals who experience mental health with the existing support system to find meaningful life so that they get a good quality of life.

The existing phenomenon figured out that people with mental disorders are increasing, and their quality of life tends to be low. Severe mental disorders in Indonesia have increased from 1.3 cases per mile to 7 cases per mile in 2018 (Ministry of Health of the Republic of Indonesia, 2018). Research also shows that the quality of life for mental disorders still tends to be low. In severe mental disorders, 67.9% have an average quality of life, 15.4% have a low quality of life, and 12.8% have a very low quality of life (Hurmuz *et al*., 2022). The quality of life of schizophrenic patients related to physical health satisfaction, physical health, psychological and social relationships, and the environment is still low (Wardani & Dewi, 2018). Another picture of the recurrence rate of mental disorders is also still high. Recurrence rates in schizophrenia of 28.0%, 43.0%, and 54.0% have been reported during the first, second, and third years respectively (Pothimas et al., 2020). Based on the concept of recovery, nursing intervention to help people with mental disorders to find the meaning of their illness is very important, not just drug-oriented.

The process of finding meaning in recovery is hindered due to factors of treatment, social support, self-assessment, belief, and spirituality, as well as physical and psychological stressors of sufferers (Gandhi, Jose, and Desai, 2020). Among these factors, spirituality is important in the journey of finding the meaning behind the illness of a person with mental health problems. In the recovery process from a severe mental disorder, spirituality becomes a predictor in helping to achieve life goals and psychological well-being so that the quality of life improves. Spirituality is strongly correlated with the recovery and psychological well-being of schizophrenic patients (Saiz *et al*., 2021a). Qualitative research on schizophrenia recovery experiences in Indonesia shows that seeking God’s help is the best way to schizophrenic life (Suryani *et al*., 2022a). Spirituality helps to find the meaning of life from the experience of illness as well as a predictor of recovery (Can Öz and Duran, 2021).

Various conceptual models in the recovery of mental disorders have been developed. It views that the recovery of mental disorders is centered on the individual being able to empower themselves with their limitations to create a meaningful life. The journey of a mental disorder to be able to control himself and have adaptive coping really needs the role of a nurse. Spirituality in nursing is a holistic concept in helping the mental disorder find the meaning of their life. Therefore the literature review is important to carry out, synthesize and identify how spirituality relates to the recovery of mental disorders. The purpose of this literature review is to analyze how spirituality affects the recovery of mental disorders.

## 2. Method

The research method used is a literature review of spirituality and recovery of mental disorders.

### 2.1 Study Protocol

The literature review guide and evaluation use a PRISMA flow diagram to determine the quality of the articles and their suitability to the theme.

### 2.2 Article search strategy

The data used in this study is the data obtained from previous studies. The literature was obtained from four reputable journal databases; Scopus, ProQuest, Science Direct, and SpringerLink.

### 2.3 Inclusion criteria

The literature studied is articles published in the last three years (2020-2022). Reviews of journals related to spirituality and mental health recovery are searched by title and abstract. Keywords used are 1) spirituality or religiosity AND recovery AND mental illness. 2) spirituality or religiosity AND recovery AND severe AND mental illness. The inclusion criteria are qualitative or quantitative research articles published in 2019-2022 in English and free access to the full text.

**Figure 1.**
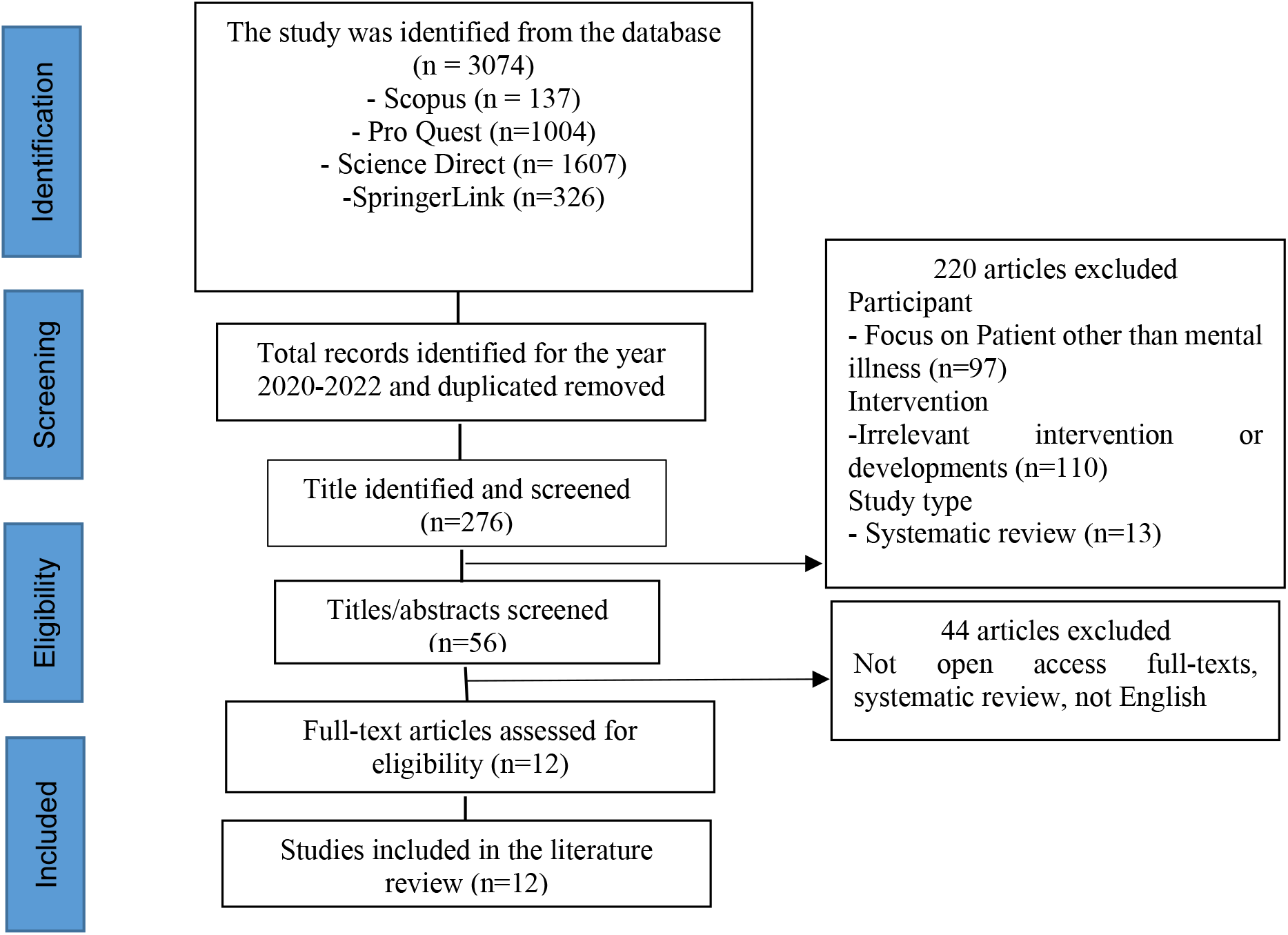
PRISMA Flowchart.

**Table 1.**
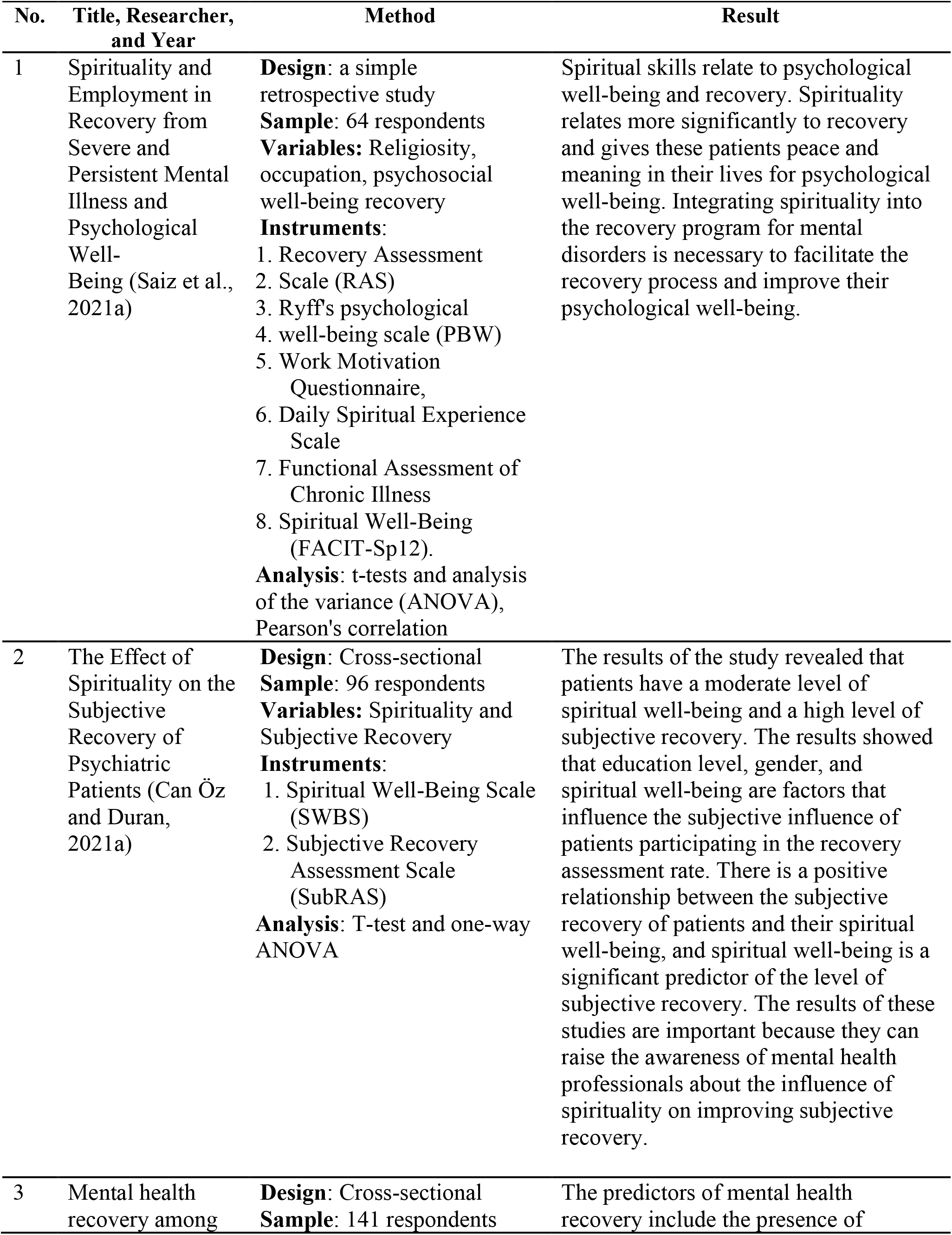
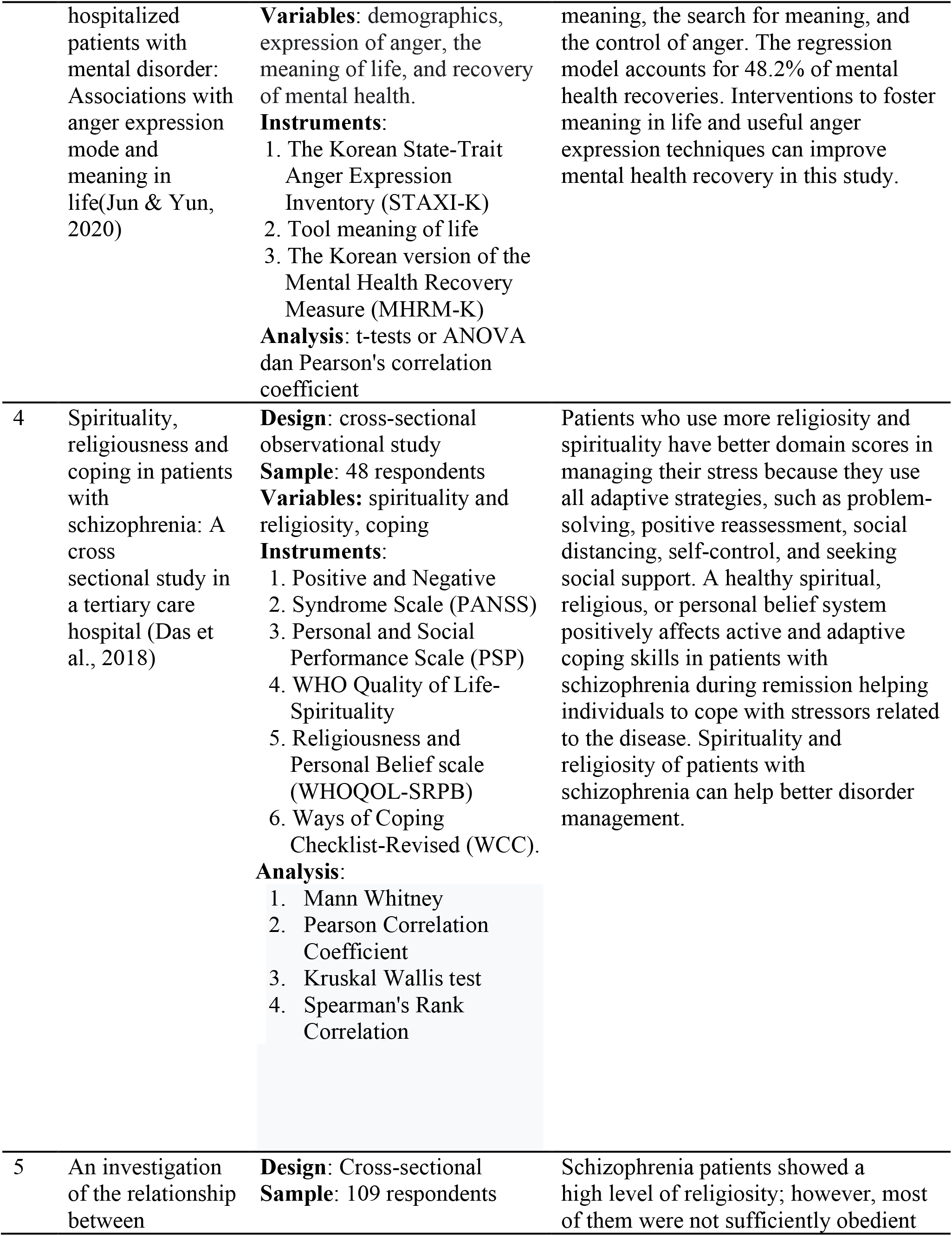
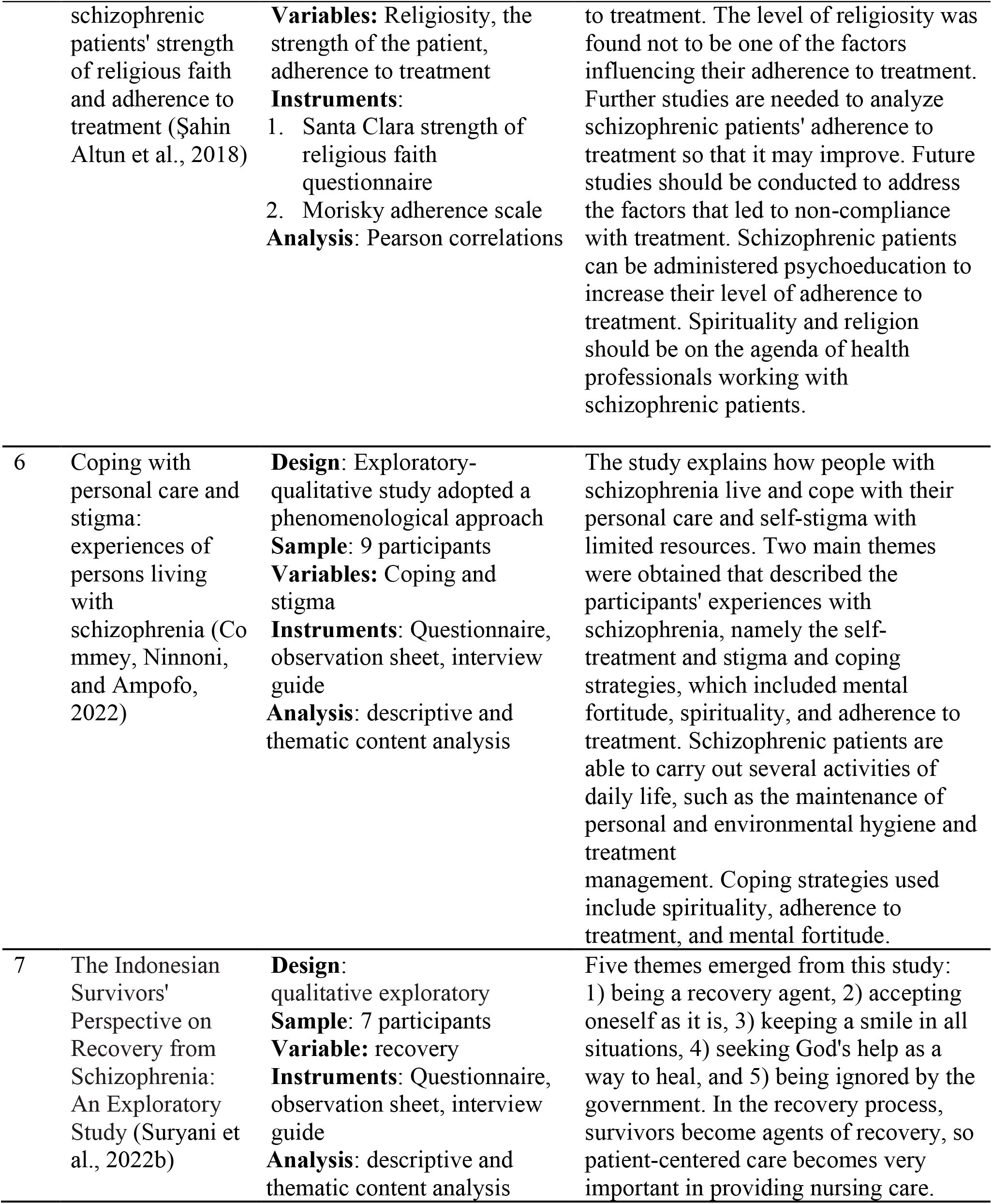
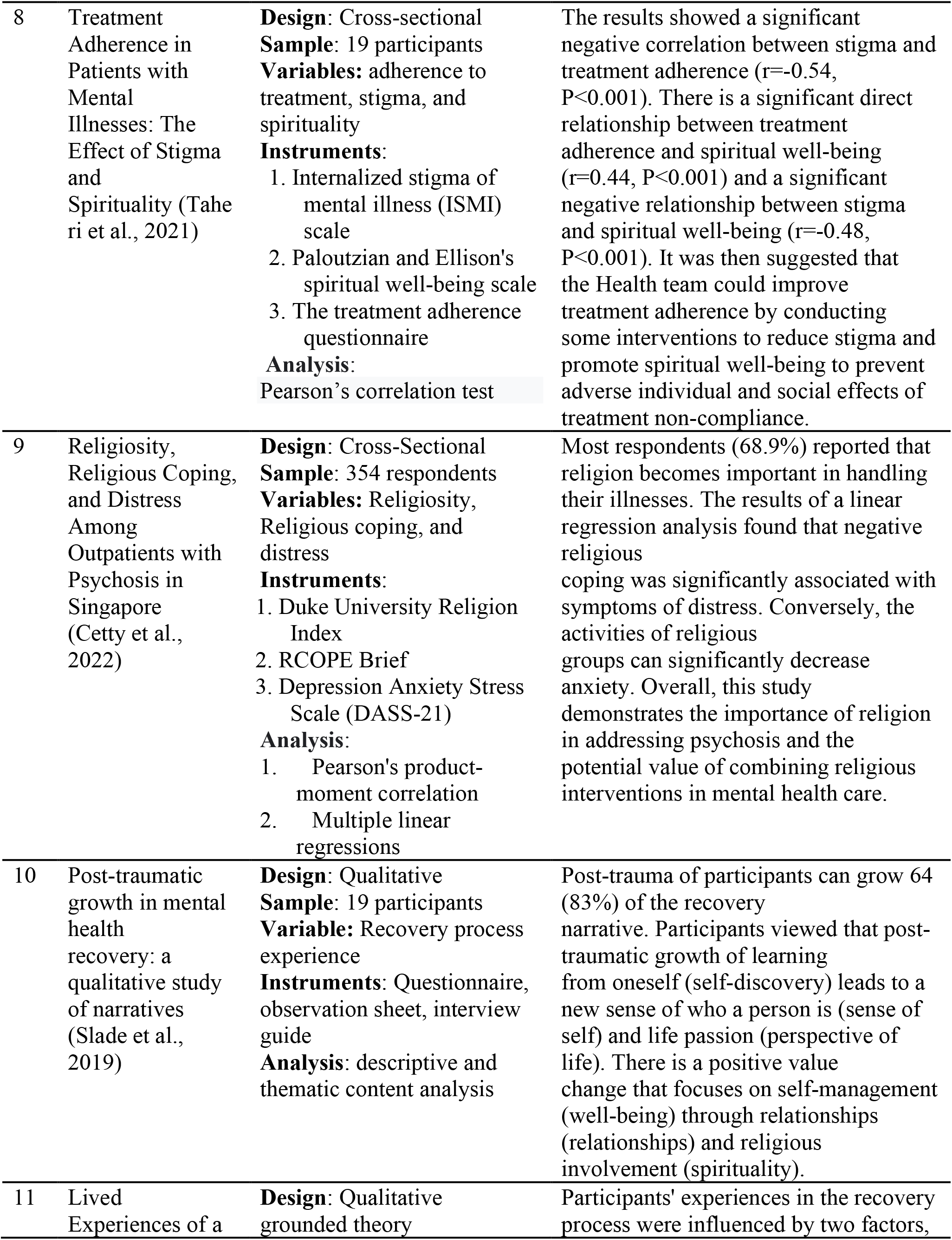
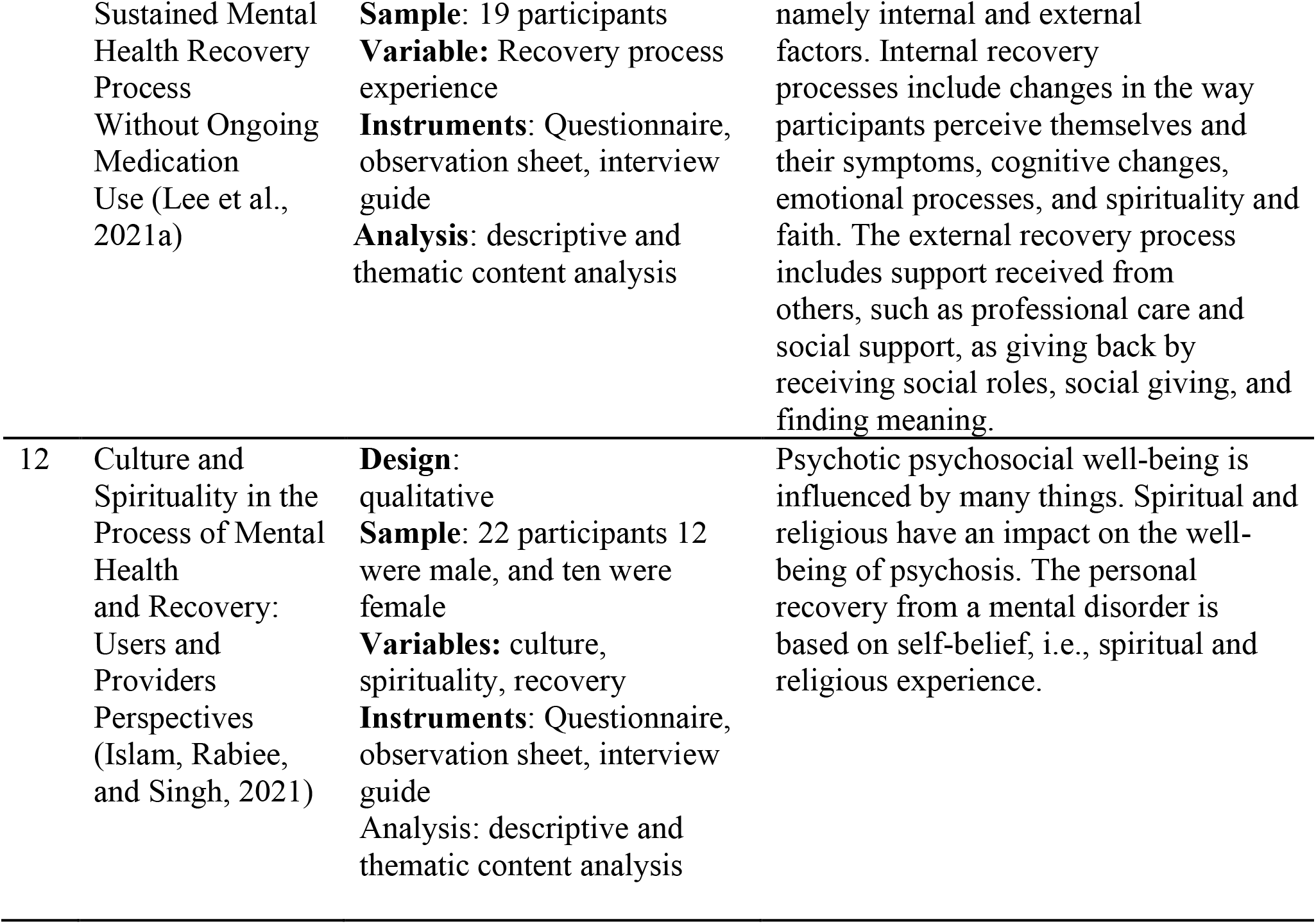
Characteristics of the literature reviewed (n=12)

## 3. Discussions

A literature review of 12 articles discusses spirituality and mental health recovery. Five qualitative research articles describe their spiritual and religious experiences as adaptive coping skills, such as 1) Finding meaning in life, 2) increasing adherence to treatment, 3) increasing self-confidence so that self-stigma decreases 4) learning from themselves about their illness. Seven articles with quantitative methods explain that spirituality and religion in mental disorders are related to recovery.

Recovery in the perspective of the Alliance Recovery Mental Health Nursing Theory views humans as someone who have the potential to grow and develop in realizing their goals. Mental health is focused on the role of mental health nurses in helping the psychological ability of people with mental disorders to improve their health. Mental health nurses assist people with mental disorders in using effective coping in exploring the abilities and resilience of sufferers and helping to increase hope, empowerment, and therapeutic relationships. (Jubb & Shanley, 2007). Hope is the basic level in the recovery of mental disorders. Hope is related to the ability to recognize and accept that there is a problem with him, commit to change through the strengths you have, plan and prioritize the future, and foster optimism (Nora Jacobson, 2001).

Another recovery component is self-empowerment which is defined as the ability to correct the inability to control themselves, a sense of powerlessness, and self-reliance. Self-empowerment includes; autonomy, namely the ability to act as an agent of self-change in terms of knowledge, self-confidence and choosing the meaning of a meaningful life; the desire to get out of feeling comfortable with the risks faced; and responsibility or a sense of responsibility for people with mental disorders to develop meaningful life through collaboration with families, groups and health workers (Nora Jacobson, 2001).

Recovering in the concept of recovery does not mean recovering to normal, but is a process of individuals with mental disorders being able to control themselves and interpret their illness. Interpreting the illness is a process of reconceptualizing the disease by facing and internalizing it as part of the life process. Self-control ability is the patient’s attempt to cope with alleviating symptoms as well as dealing with psychological and social problems. The self-control strategy in recovery concept for a person with mental disorders is the ability to be an agent and control in controlling his or her life (Nora Jacobson, 2001).

Spirituality and faith help people with mental disorders find meaning in their lives (Lee et al., 2021; Saiz et al., 2021). Spirituality is the tendency to make meaning through one’s interpersonal relationships, interpersonal with transpersonal, which empowers a person to overcome problems. Spirituality is a cultural factor that provides structure and human values, behavior, and experiences (Ah. Yusuf, 2017). The middle-range theory of spiritual well-being in illness is used as a holistic nurse intervention using the body, mind, and spiritual perspectives. This theory is an integration of the theory of spiritual well-being and a conservative nursing model where the main focus of the theoretical framework is to find meaning from life experiences during illness. The main concept of this theory is that sick individuals are helped to be able to find the experience of spiritual meaning in their illness to gain inner well-being. (O’Brien, 2011). Therefore nurses helping the recovery of mental health in mental disorders by assisting the client’s spiritual coping skills is an effort to carry out self-responsibility to get the meaning of his life.

Mental health nurses help people with mental disorders through religious and spiritual attitudes and behaviors in interpreting their illness. A descriptive, cross-sectional study conducted by patients in a psychiatric clinic at a hospital stated that spiritual well-being is one of the factors influencing recovery. Spiritual well-being is positively related to recovery, and spiritual well-being is a significant predictor of the recovery rate of people with mental disorders. Therefore, psychiatric nurses are expected to focus on serving patients through a holistic healthcare approach and increasing the patient’s subjective recovery rate through interventions that will strengthen the spiritual dimension (Can z & Duran, 2021). Other studies have found helping patients find meaning in their lives to be a more significant element in their recovery as well as for psychological well-being. Integrating spirituality into recovery programs for people with mental disorders is necessary to facilitate the recovery process and improve their psychological well-being (Saiz et al., 2021).

In the context of recovery, the strength of the nurse-patient relationship to be able to raise the strengths they have, build hope and find meaning in life is an essential element. Nurses must be able to apply humanistic principles and avoid stigma against patients. Partnership relationships between nurses and patients are carried out to help build hope, foster self-responsibility, explore strengths and help find meaning. The stages of intervention that nurses can do include (Jubb & Shanley, 2007):

1. The first stage is establishing a partnership relationship between nurses and mentally ill people to form values.
2. The second stage is that the nurse takes responsibility for identifying and prioritizing the problem and setting goals with the sufferer.
3. The third stage is that nurses and mentally ill people empower to apply recovery principles.
4. Stage four is helping people with mental disorders use their own coping strategies to achieve their goals of accepting self-responsibility and controlling their well-being.

Therefore, the ability of the nurse who is prepared to carry out recovery interventions in mental patients includes the following:

1. The psychiatric nurse appreciates every communication that the sufferer conveys to build the strength of the relationship.
2. The psychiatric nurse always provides time to help the sufferer whenever necessary
3. The psychiatric nurse is constantly open in therapeutic communication to help the recovery
4. Psychiatric nurses must have therapeutic communication skills so that the barrier to a negative assessment of the sufferer can be eliminated
5. The psychiatric nurse must be able to view the holistic philosophy of the mental disorder patients by appreciating every experience they have to help find the meaning of their life.

## 4. Conclusion

Psychological management based on spirituality in mental disorders can improve coping skills to build self-confidence, raise strengths and be able to find the meaning of life. Self-confidence and finding the meaning of life in a mental disorder is a form of psychological well-being that improves their quality of life. Spiritual interventions integrated with mental health nursing are essential to improve the recovery of mental disorders.

## Data Availability

All data produced in the present work are contained in the manuscript

## Notes

### Competing Interest Statement

The authors have declared no competing interest.

### Funding Statement

This study did not receive any funding

### Author Declarations

The study used ONLY openly available human data that were originally located in the database publication (Scopus, Proquest, Science Direct and SpringerLink)

